# Hyaluronic acid for tissue and bone regeneration after tooth extraction. Systematic review and meta-analysis

**DOI:** 10.1101/2020.06.02.20120733

**Authors:** Shuheng Lai, Francisco Novillo, Geovanna Cárdenas, Francisca Verdugo, Gabriel Rada

**Affiliations:** Faculty of Medicine, Pontificia Universidad Católica de Chile, Santiago, Chile; Fundación Epistemonikos, Santiago, Chile; UC Evidence Center, Cochrane Chile Associated Center, Pontificia Universidad Católica de Chile, Santiago, Chile; Internal Medicine Department, Faculty of Medicine, Pontificia Universidad Católica de Chile, Santiago, Chile

## Abstract

**Objective:** The objective of this systematic review is to assess the effectiveness and safety of Hyaluronic Acid (HA) on tissue and bone regeneration after tooth extraction.

**Data Sources:** We will conduct a comprehensive search in Epistemonikos, PubMed/Medline, Embase, Cochrane Central Register of Controlled Trials (CENTRAL), LILACS, the International Clinical Trials Registry Platform (ICTRP), ClinicalTrials.gov, US National Institutes of Health (NIH) and grey literature, to identify all relevant randomized controlled trials regardless of language or publication status (published, unpublished, in press and in progress).

**Eligibility Criteria for Selecting Studies and Methods:** We will include randomized trials evaluating the effect of HA on tissue and bone regeneration after tooth extraction. Two reviewers will independently screen each study for eligibility, data extraction, and assess the risk of bias. We will pool the results using meta-analysis and will apply the GRADE system to assess the certainty of the evidence for each outcome.

**Ethics and Dissemination:** No ethics approval is considered necessary. The results of this review will be disseminated via peer-reviewed publications, social networks, and traditional media.

**PROSPERO Registration ID:** CRD42020150285

## Introduction

A dental extraction is the surgical procedure of removing a tooth, either primary or permanent, performed by a dentist (1). The main causes reported for dental extractions are caries for adults aged 50 years or younger, orthodontic related motives in children, whilst periodontal disease is the main cause in the elderly (2).

After dental extraction, the remaining tooth socket is left without periodontium, therefore, the stimuli that maintain bony level is lost, causing an irreversible and progressive resorption of the alveolar bone (3). This bone loss diminishes in turn, the supporting tissue altering the appearance of the patient causing speech and masticatory impairment and makes an adequate restoration harder to achieve (4).

In order to prevent some of these negative effects of bone resorption, different biomaterials can be used as an adjunctive therapy to promote tissue repair and bone regeneration in the tooth socket after the procedure, lower the alveolar bone resorption rate (5) (6). These compounds can be of biological, synthetic, or composite material origin (7) (8).

Hyaluronic acid (HA) is a glycosaminoglycan (GAG) with a high molecular weight. HA is one of the main components of the extracellular matrix and is widely distributed in different tissues (9).

HA has been recently studied as a topical adjunctive therapy in chronic inflammatory conditions, as well as an aid in tissue repair for dental procedures. Depending on the use case, different formulations and concentrations have been tested (10). Several clinical trials have shown a beneficial effect of topical application of HA over tissue repair and regeneration, this evidence suggests that HA application in oral surgery could have a significant role in improving post-surgical outcomes (11) (12) (13).

Despite being widely used in different medical fields as an anti-inflammatory agent, HA’s application in dentistry have not been completely explored (10). There is one review evaluating the use of HA as a biomaterial applied after dental extractions (14).

The currently available evidence on this subject is inconclusive. The aim of this systematic review is to provide a rigorous up to date summary of the actual evidence on the efficacy and safety of topically applied HA in tissue repair and bone regeneration, on tooth sockets after dental extraction.

## Methods

### Types of studies

We will include randomized controlled trials (RCTs) meeting our inclusion criteria and reporting useable data, with no restriction on their length of follow-up. Quasi-randomized trials (QRTs) will not be included. We will exclude studies evaluating the effects on animal models or in vitro conditions.

### Types of participants

We will include trials assessing healthy individuals aged 12 and above, with complete permanent dentition who underwent a surgical tooth extraction for any cause.

### Types of interventions

We will include studies investigating the use of HA (any formulations), over the surgical site after a dental extraction compared to placebo, no treatment or another active therapy.

Trials that employ HA as an adjunctive therapy to treat conditions other than a dental extraction or consider other active treatments in conjunction with HA will be excluded.

## Types of outcome measures

We will not use outcomes as inclusion criteria, during the selection process. Any article meeting all the criteria except for the outcome criterion will be preliminarily included and assessed in full text.

### Primary outcomes

Pain, swelling, trismus, postsurgical bleeding, site infection and adverse effects.

### Secondary outcomes

Time to complete tissue repair and bone regeneration.

## Search methods for identification of studies

### Electronic searches

We will conduct a comprehensive search in PubMed/Medline, Embase, Cochrane Central Register of Controlled Trials (CENTRAL), Lilacs, the International Clinical Trials Registry Platform (ICTRP), ClinicalTrials.gov, US National Institutes of Health (NIH) and grey literature, to identify all relevant randomized controlled trials regardless of language or publication status (published, unpublished, in press and in progress). The searches will cover from the inception date of each database until the day before submission.

The following search strategy will be used to search in Pubmed/Medline. We will adapt it to the syntax of other databases.

#1 Tooth Extraction[MeSH Terms]; #2 Tooth Socket[MeSH Terms]; #3 ((dental* OR tooth* OR teeth*) AND (socket OR extraction* OR missing OR loss OR lost)); #4 exodonti*; #5 #1 OR #2 OR #3 OR $4; #6 Hyaluronic Acid[MeSH Terms]; #7 (hyaluronic acid); #8 hyaluronan; #9 hyaluronic*; #10 hyaluronate; #11 “sodium hyaluronate”; #12 (Gengigel OR Hyadent OR Hylodent OR Oddent); #13 #6 OR #7 OR #8 OR #9 OR #10 OR #11 OR #12; #14 randomized controlled trial [pt]; #15 controlled clinical trial [pt]; #16 randomized [tiab]; #17 placebo [tiab]; #18 drug therapy [sh]; #19 randomly [tiab]; #20 trial [tiab]; #21 groups [tiab]; #22 #14 OR #15 OR #16 OR #17 OR #18 OR #19 OR #20 OR #21; #23 (animals [mh] NOT humans [mh]); #24 #22 NOT #23; #25 #5 AND #13 AND #24

### Searching other resources

An expanded search will be performed to identify articles potentially missed through the database searches and in order to identify ‘grey literature’ and unpublished studies. This includes the following:

- MEDLINE for systematic reviews addressing the same question as our review.
- In order to find additional literature, we will run a Google Scholar search for key terms and authors.
- We will hand search reference lists of all included studies and of relevant reviews retrieved by the electronic searching to identify further relevant trial.
- Authors of included studies will be contacted for any additional published or unpublished data.

### Selection of studies

The results of the literature search will be uploaded to the screening software CollaboratronTM (15). In CollaboratronTM (15), two researchers will independently screen the titles and abstracts yielded by the search against the inclusion criteria. We will obtain full-text reports for all potentially eligible studies that appear to meet the inclusion criteria or require further evaluation to decide about their inclusion. The same review authors will assess independently those articles and decide on fulfilment of inclusion criteria. A third review author will resolve discrepancies in the case of disagreement. Articles retrieved from the screening and included in the review will be recorded in RevMan 5.3 (16). Excluded trials after full text revision and the primary reason for the decision will be listed.

The selection process will be documented in a in a PRISMA flow diagram (17) adapted for the purpose of this project.

### Extraction and management of data

Using standardized forms, two reviewers will independently extract data from each included study. We will collect the following information: study design, setting, baseline characteristics of patients and eligibility criteria; details of intervention, co-interventions and comparison; number of patients assigned to each treatment group, the outcomes assessed and the time they were measured; number of patients with adverse reactions per treatment group and method used to seek adverse reactions. Losses to follow up, exclusions, and the reasons accordingly. A third review author will resolve discrepancies in the case of disagreement.

### Risk of bias assessment

Risk of bias in included studies will be assessed according to the ‘Risk of bias’ table, which is the tool recommended by The Cochrane Collaboration (18) (19). Descriptions and judgements about the following domains will be included: adequacy of sequence generation, allocation concealment, blinding, addressing of incomplete outcome data, likelihood of selective outcome reporting, and other potential sources of bias.

Risk of bias table domains will be classified as “low risk” “high risk” or “unclear risk”. The ‘Risk of bias’ table will be filled independently by two review authors, with a third review author acting as arbiter.

Original study authors will be contacted for further information if one of the review authors requires clarification wherever necessary.

### Measures of treatment effect

Pooled dichotomous outcomes will be reported as risk ratios (RR) or odds ratios (OR) with 95% confidence intervals (CI), and continuous outcomes will be reported as mean difference (MD) with 95% CI. Outcomes using different scales (for example, different measurements of facial edema) will be reported as standardized mean differences (SMD) with a 95% CI.

Then, these results will be displayed on the ‘Summary of Findings Table’ as mean difference.

### Dealing with missing data

Main author of trials will be contacted in order to verify key study characteristics and obtain missing numerical outcome data. If not possible, only available data will be analyzed. The potential impact of missing data will be addressed in the ‘[Discussion](https://docs.google.com/document/d/1JXtcUhAJ3cBMgmg8dP1_ScURXY1V-oTEg-op6msJuxM/DISCUSSION)’ section, analysis of worst case scenario and best case scenario will be applied.

If no intention-to-treat analysis (ITT) is reported, or the ITT analysis has been modified, data will be reanalyzed following intention to treat principles if possible. If not possible, this will be addressed in the ‘[Discussion](https://docs.google.com/document/d/1JXtcUhAJ3cBMgmg8dP1_ScURXY1V-oTEg-op6msJuxM/DISCUSSION)’ section.

### Assessment of heterogeneity

Heterogeneity will be quantitatively assessed with a statistical test (Q statistic) and the I^2^ statistic. Statistically significant heterogeneity will be defined as at least one positive test (establishing a cut-off value of P = 0.10 for the Mantel-Haenszel Chi^2^ test, or values over 50% using the I^2^ statistic).

### Assessment of reporting biases

Publication bias will be evaluated through visual analysis of funnel plots. Evidence of asymmetry will be based on P < 0.10, and present intercepts with 90% CIs. Discrepancies between registered protocols and final publications will be assessed to evaluate other reporting biases including outcome reporting bias. If no record is found for a study in the WHO International Clinical Trials Registry Platform, authors will be contacted for further information.

### Data synthesis

Statistical analysis will be performed in accordance with the guidelines for statistical analysis developed by Cochrane. If applicable, studies will be pooled to perform meta-analysis comparing HA application versus placebo or other treatment for each outcome measure. When possible, meta-analyses using the random-effects inverse variance model will be carried out to estimate the pooled measure of treatment effect, fixed effects models will be performed to evaluate if findings are not sensitive to choice of analysis. If specific populations or interventions are found to be significantly different, separate meta-analyses will be performed to assess if heterogeneity is explained by some of these, or if a convincing subgroup effect is found. If discrepancies between analyses are found, evidence will be presented and addressed in the ‘[Discussion](https://docs.google.com/document/d/1JXtcUhAJ3cBMgmg8dP1_ScURXY1V-oTEg-op6msJuxM/DISCUSSION)’ section.

### Subgroup analysis and investigation of heterogeneity

If enough data is available, subgroup analysis will be performed to assess any outcome differences depending on formulation, concentration, and administration of HA.

There is no standard application, dosage or formulation for HA, as adjunctive therapy for dental extraction, therefore, it is relevant to classify and analyze these variables, as they might affect the outcomes.

### Sensitivity analysis

Sensitivity analyses will be performed to evaluate the impact of the inclusion or exclusion of missing data and the choice of a fixed-effect or random-effects models. If there are any substantial differences, data from lower-quality studies will not be pooled with the higher-quality studies, and will be presented in different analyses, unless separate analyses do not differ greatly from pooled results.

### Summary of Findings

The certainty of the evidence for all outcomes will be reviewed using the Grading of Recommendations Assessment, Development and Evaluation working group methodology (GRADE Working Group) (20). Findings for the main outcomes will be summarized in Summary of Findings (SoF) tables.

## Data Availability

All data generated or analysed during this study are included in this published article (and its supplementary information files).

## Notes

### Roles and contributions

SL conceived the protocol, SL, FN and GC drafted the manuscript, and all other authors contributed to it. The corresponding author is the guarantor and declares that all authors meet authorship criteria and that no other authors meeting the criteria have been omitted.

### Ethics

As researchers will not access information that could lead to the identification of an individual participant, obtaining ethical approval was waived.

### PROSPERO registration

This protocol has been submitted PROSPERO Registration ID CRD42020150285.

